# A Framework for Automated Gene Selection in Genomic Screening

**DOI:** 10.1101/2020.12.11.20231449

**Authors:** L Lazo de la Vega, W Yu, K Machini, CA Austin-Tse, L Hao, CL Blout Zawatsky, H Mason-Suares, RC Green, HL Rehm, MS Lebo

**Affiliations:** Laboratory for Molecular Medicine, Mass General Brigham Personalized Medicine, Cambridge, MA; Department of Pathology, Brigham & Women’s Hospital, Boston, MA; Harvard Medical School, Boston, MA; Medical and Population Genetics, The Broad Institute of MIT and Harvard, Cambridge, MA; Center for Genomic Medicine and Departments of Pathology and Medicine, Massachusetts General Hospital, Boston, MA; Division of Genetics, Department of Medicine, Brigham and Women’s Hospital, Boston, MA, USA; Ariadne Labs, Boston, MA

**Keywords:** Clinical genetic testing, gene-disease association, genomic screening, gene curation

## Abstract

An efficient framework to identify disease-causing genes is needed to evaluate genomic data for both individuals with an unknown disease etiology and those undergoing genomic screening. Here, we propose a framework for gene selection used in genomic analyses, including screening applications limited to genes with strong or established evidence levels and diagnostic applications that includes genes with less or emerging evidence of disease association. We extracted genes with evidence for gene-disease association from the Human Gene Mutation Database, Online Mendelian Inheritance in Man, and ClinVar to build a diagnostic gene list of 5,973 genes. Next, we applied stringent filters in conjunction with computationally curated evidence (DisGeNET) to create a list limited to 3,600 genes with stronger levels of evidence for disease association. When compared to manual gene curation efforts, including the Clinical Genome Resource, genes with strong or definitive disease associations are included in both gene lists at high percentages, while genes with limited evidence are largely removed. We further confirmed the utility of this approach in the screening of 45 ostensibly healthy genomes. Our approach efficiently creates highly sensitive gene lists for genomic applications, while remaining dynamic and updatable, enabling time savings in gene curation and review.

## INTRODUCTION

As genome and exome sequencing become standard in clinical genetic testing for patients with unknown genetic etiology and in broad genomic screening for population precision health, an efficient framework to identify and capture all known disease-causing genes and variants is needed. With the scope of analysis in these assays covering over 20,000 genes, it is challenging to rapidly determine which genes have evidence of clinical relevance. To limit the interpretative burden of reviewing variants from all genes, a well-defined “medical exome” is needed, consisting of genes with sufficient levels of evidence to warrant review in a clinical assay.

There have been efforts to establish highly curated lists of gene-disease associations (GDAs), but these are often small, though highly curated. Most notably, the Clinical Genome Resource (ClinGen) has established a robust framework to determine gene-disease validity through manual assessment of strength of evidence that is used within their multiple disease-specific expert panels and working groups^1^. While these GDAs are well-curated, the intense effort required has limited the breadth of genes currently annotated. On the other end of the spectrum, computational tools, such as DisGeNET, attempt to classify the GDAs of all genes by integrating multiple databases into a single GDA score^2-4^. However, the accuracy and validity of this scoring system has not been assessed. Other efforts have taken the approach of crowd-sourcing the GDAs, such as Genomics England’s PanelApp, which allows diagnostic gene panels to be shared, downloaded, and evaluated by the scientific community, though they may be limited by the interests and thoroughness of the submitters^5^.

Generating and maintaining up-to-date gene lists remains challenging since assessing all GDAs would be prohibitively time-consuming and evidence supporting new and existing GDAs is continuously generated. Previously published projects from our group, BabySeq and MedSeq, required manual curation resulting in a list of 1,514 and 1,490 GDAs, respectively. In both projects, this was a labor intensive and time-consuming process that is not easily replicated in an efficient manner^6, 7^. Therefore, a balance between efficiency and thoroughness is required to make the analysis of genomic data more feasible.

Here, we propose a framework to create gene lists for genomic analyses that balances efficiency, robustness, and accuracy with the ability to be routinely updated with new genes as associations emerge from the literature. This approach generates two lists of disease-associated genes based on different levels of evidence to be used in diagnostic assays and genomic screens.

## METHODS

### Data Sources Used to Generate the Diagnostic and Screening Gene Lists

Comprehensive databases of gene and/or variant associations, including the Human Gene Mutation Database (HGMD), ClinVar, Online Mendelian Inheritance in Man (OMIM), and DisGeNET, were used to identify genes with any reported GDA ^2-4, 8-10^. Each data source was specifically parsed to identify, when applicable, the number of classified variants and their review date, publications, and gene identifiers **(Supplemental Methods)**.

### Data Sources Used for Validation of Diagnostic and Screening Gene Lists

Data sources incorporated for gene list validation included: 1) 1,490 GDAs evaluated in MedSeq^7^; 2) 1,514 GDAs evaluated in BabySeq^6^; 3) 1,043 gene curations in 824 genes captured by ClinGen as of August 12, 2020^1^; and 4) 232 and 306 panels from the PanelApp Australia and Genomics England PanelApp, respectively (accessed July 23, 2020)^5^. Each dataset included a list of GDAs and their strength of evidence. Classifications used in each dataset and how they map to an overall strength of evidence are provided in **Table S1**, and defined in **Supplementary Methods**.

### Genome Sequencing and Analysis

Genome sequencing data was generated from 45 individuals undergoing non-indication based genomic screening **(Supplemental Methods)**, with >30X mean coverage and a minimum completeness of >95% of all bases at 15X or higher. Variants were filtered to the diagnostic or screening gene lists to identify Pathogenic (P) or Likely Pathogenic (LP) variants **(Supplementary Methods)**. Only genes mapping to GRCh37 were analyzed **(Table S2)**. Evidence for GDAs were manually curated and each GDA was assigned one the following categories: (1) Definitive, (2) Strong, (3) Moderate, or (4) Limited using ClinGen criteria for gene-disease association. Following gene and variant curation using 2015 American College of Medical Genetics and Genomics/Association of Molecular Pathology guidelines ^11^ with ClinGen rule specifications, only P/LP variants in genes with a strong or definitive GDA were considered reportable. The filtered and reportable variants were evaluated among this set of 45 genomes for the diagnostic and screening gene list framework described here.

## RESULTS

### Generated Diagnostic and Screening Gene List

To build a comprehensive diagnostic gene list, we extracted all genes from the 3 downloaded datasets that meet any of the following criteria: 1) ≥1 P/LP variant in ClinVar, excluding CNVs overlapping multiple genes; 2) ≥1 variant classified as a disease-causing mutation (DM) in the HGMD; and 3) listed in Morbid Map from OMIM, excluding susceptibility and non-disease genes **(Figure 1A)**. Following these filters, the diagnostic exome list included 5,973 genes that have been implicated in Mendelian disease that can be used in a diagnostic setting. Of note, 3,741 genes were present in all 3 datasets, with HGMD contributing the most unique genes **(Figure 1B)**.

**Figure 1:**
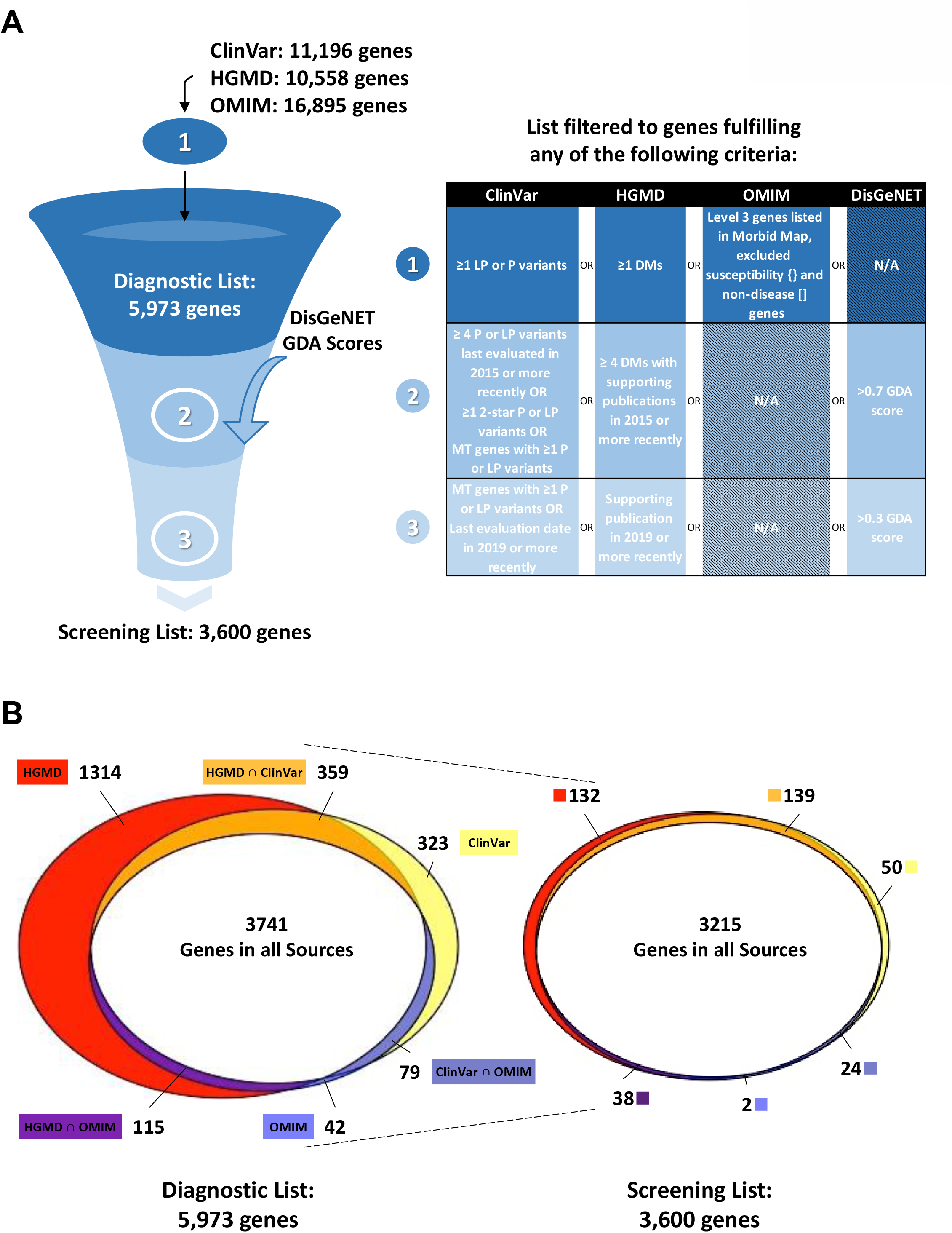
(A) Schematic of the criteria fulfilled at each stage of the gene filtration process. Genes with entries in ClinVar (11,196 genes), OMIM Morbid Map (16,895 genes), and HGMD (10,558 genes) were integrated to generate the diagnostic and screening gene list. Filtration parameters for each stage are presented in the right panel. (B) Venn diagram of the diagnostic (left) and screening (right) gene list, including the number of genes meeting criteria in the initial databases.

For genomic screening applications, where reportable variants are often limited to genes with stronger disease associations, the diagnostic gene list was further filtered to keep genes that fulfilled any of the following criteria: 1) ≥4 P/LP variants in ClinVar evaluated in 2015 or more recently by any submitter; 2) ≥1 2-star P/LP variant in ClinVar; 3) mitochondrial genes with ≥1 P/LP variant in ClinVar; 4) ≥4 DMs in HGMD with supporting publications in 2015 or more recently; 5) genes with a DisGeNET GDA score >0.7. To add more stringency, we filtered this intermediate list to remove genes with lower levels of evidence, only keeping genes that met at least one of the following criteria 1) ≥1 DM in HGMD with a supporting publication in 2019 or more recently, 2) ≥1 P/LP variant with a last evaluated date in ClinVar in 2019 or more recently, or 3) genes with a DisGeNET GDA score >0.3. All mitochondrial genes already in the filtered list were also kept at this stage. After applying these additional filters, a screening gene list of 3,600 genes remained, with 3,215 genes present in all original data sources **(Figure 1)**.

### Comparing Gene Lists to Previous Curations

To determine the utility of the gene lists and specificity of the filtering strategy, we compared the diagnostic and screening list to various manual approaches, including rigorous curations by experts in ClinGen, manual assessments by an individual lab for BabySeq and MedSeq, and crowd-sourced approaches in PanelApp. When both lists were compared to the 824 genes from ClinGen, we observed that all gene-disease pairs classified as definitive (538 genes) or strong (19 genes) by ClinGen were captured by both lists except for two definitive GDAs missing from the screening list: *F13B* associated with Factor XIIIB deficiency and *GP6* associated with Platelet-type bleeding disorder 11. Neither were captured by the screening list as each only had 3 P/LP variants in ClinVar and no publications after 2014 in HGMD **(Table S2)**. Some gene-disease pairs with limited, disputed, refuted, or no evidence were removed from the diagnostic list (4.6%; 8/173) and many of these genes were removed from the screening list (34.7%; 60/173) **(Figure 2A)**.

**Figure 2:**
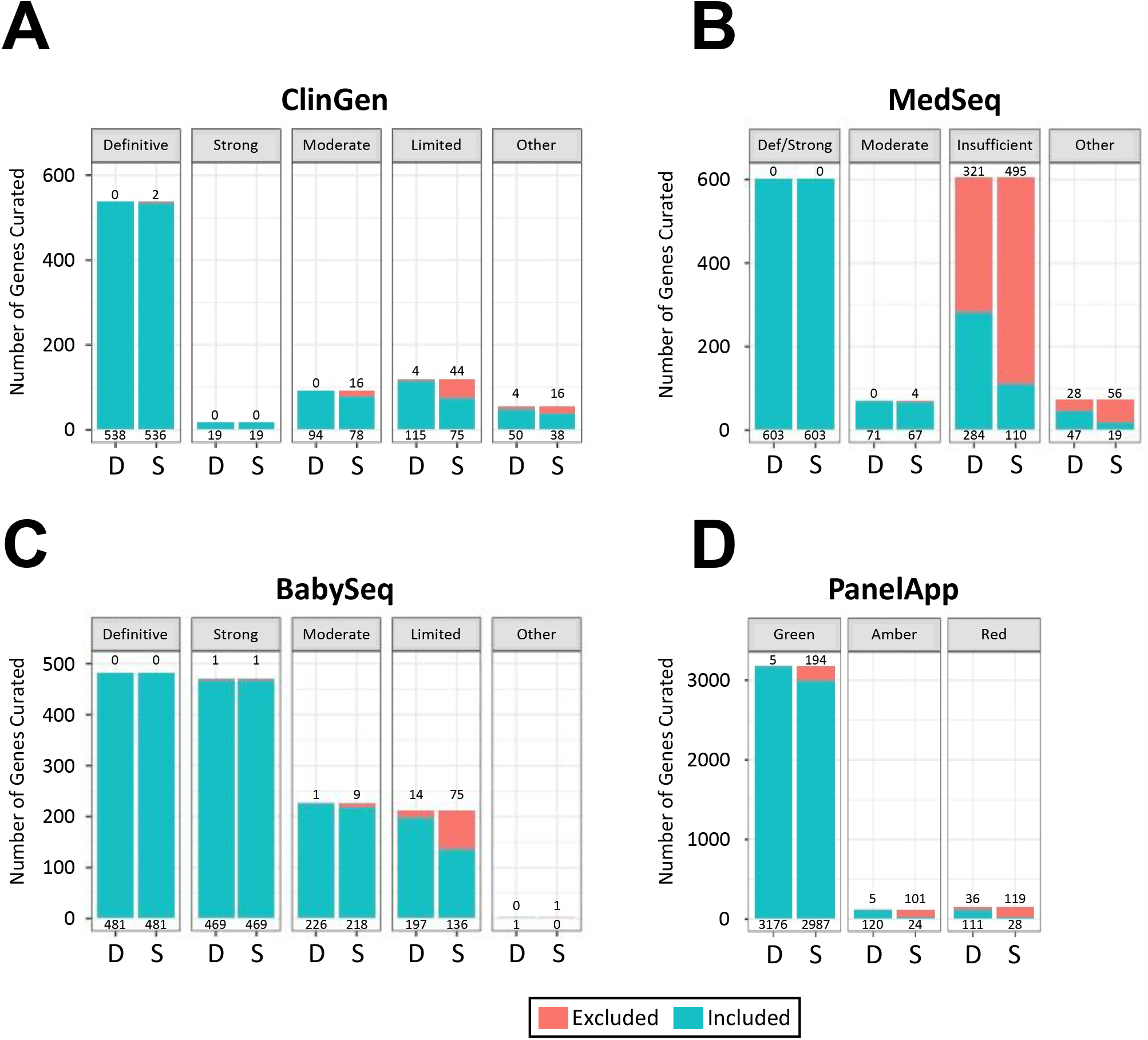
Diagnostic and screening gene lists were compared to the GDA classifications assigned by 4 resources (A) ClinGen, (B) MedSeq, (D) BabySeq and (C) intersection of PanelApp Australia and Genomics England PanelApp. Numbers below the bar represent the number of genes included and numbers above the bar are the number of genes excluded in the respective list. Other: conflicting, refuted, disputed, no reported evidence, trait, pharmacogenomic association, only claim is from GWAS, and does not meet criteria; D: Diagnostic Gene List; S: Screening Gene List.

Comparing the gene lists to the more rapid assessments across a broad range of genes in MedSeq or BabySeq^6, 7^, we observed that all gene-disease pairs classified as definitive or strong in both studies (603 genes and 951 genes, respectively) were captured by both lists, except for the strong *RPS15* association with Diamond-Blackfan anemia curated in BabySeq that was not included in either gene list. The GDA between *RPS15* and Diamond-Blackfan anemia was reassessed by the BabySeq team and downgraded to limited due to lack of supporting evidence. The diagnostic and screening gene lists also removed 51.3% (349/680) and 81.0% (551/680), respectively, of genes with insufficient or other classifications reported in MedSeq **(Figure 2B)** and 6.6% (14/212) and 35.8% (76/212), respectively, of genes with a limited or other classification as reported in BabySeq **(Figure 2C)**.

An additional analysis was performed using gene curations reported in 534 panels generated by the PanelApp Australia and Genomics England PanelApp^5^, which rate genes according to level of evidence using a traffic-light system; green represents high-level, amber represents moderate-level and red represents low-level of evidence. Most green-rated genes (3,181 genes) were captured by both lists except for 5 and 194 genes which were removed from the diagnostic and screening gene list, respectively. The relatively high number of green-rated genes excluded from PanelApp in our screening list is expected as PanelApp is primarily focused on providing gene panels to be used in a diagnostic rather than screening setting, and have not necessarily undergone extensive GDAs using rigorous criteria, such as is used in ClinGen. The gene lists also removed 24.5% (36/147) and 81.0% (119/147) of red genes from the diagnostic list and screening list, respectively **(Figure 2D)**.

### Genome Sequencing Results Using Different Gene Lists

To determine the performance of the gene lists in case analysis, 45 genomic data sets from 45 individuals were screened for reportable variants using both the diagnostic and screening gene lists. Following variant filtration for putative P/LP variants across all 45 individuals, a total of 1,374 variants were identified within the diagnostic gene list; only 1,135 were present using the screening list, a removal of 239 variants (17%**; Figure S1A)**. Per individual, this equated to an average of 31 (min=14; max=52) and 25 (min=13; max=46) variants in the diagnostic and screening gene lists, respectively. While 57% (401/706) of the genes in the diagnostic list met Strong or Definitive disease association after manual review, this ratio increased to 74% in the screening list (401/544). After manual review of the variants in both lists, all reportable variants from the larger diagnostic list – defined as P/LP associated to a strong or definitive GDA – were also identified in the screening gene list (an average of 3 variants per individual; min=0; max=7) **(Figure S1B)**.

## DISCUSSION

Part of an effective and efficient strategy for exome and genome analyses includes defining an appropriate list of genes to interrogate for disease-causing variants. All genes with evidence for a disease association are needed for expanded analyses. However, in different contexts, the level of evidence required for the GDA may vary. For instance, genes with less or emerging evidence of disease association may be useful in a diagnostic setting where additional familial studies can help determine the likelihood of the gene’s responsibility for the individual’s disease. However, lists including limited evidence genes will have less utility in the context of genomic screening where the asymptomatic individual will not contribute evidence to the GDA and there is no or very limited utility of returning the result.

Here, we provide a framework that utilizes available databases to efficiently generate both a diagnostic list (5,973 genes) and a screening list (3,600 genes) **(Figure 1; Table S1)**. The more stringent screening gene list excluded a large percentage of genes (34.7-81%) with lower levels of evidence based upon manually curated datasets, while maintaining all strong or definitively associated genes, aside from two genes with older and borderline levels of evidence **(Figure 2)**. Additionally, using the screening gene list in 45 healthy genomes captured all reportable variants that were found using the diagnostic list.

Further refinements to this approach can help reduce the burden of comprehensive genomic screens even further, including utilizing more variant level information in the approach, such as handling variants with discordant classifications and variants whose population frequencies may suggest they may be too common to be associated with Mendelian disease. However, our current approach is easily implemented and updatable, shows high performance when compared to manually curated datasets, and can provide increased efficiency as genomic applications become more routine.

## Data Availability

The resulting genes lists and information used to create them can be found in the Supplemental Materials. This gene list will be provided on-line for easy access and on-going updates.

https://docs.google.com/spreadsheets/d/1VabFne_4TqEHxczwurwC8gokE0Q9fXCfbLlS5-9SqYs/edit#gid=1463456679

## Notes

### Competing Interest Statement

Ms. Blout Zawatsky, Dr. Lazo de la Vega, Dr. Lebo, report grants from National Heart Blood and Lung Institute, during the conduct of the study. Dr. Austin-Tse, Dr. Mason-Suares, Dr. Machini, Dr. Hao, and Dr. Yu have nothing to disclose. Dr. Rehm reports grants from NIH, grants from National Heart Blood and Lung Institute, during the conduct of the study; personal fees from Genome Medical, outside the submitted work. Dr. Green reports grants from National Heart Blood and Lung Institute, during the conduct of the study; personal fees from AIA, personal fees from SavvySherpa, personal fees from Verily, personal fees from Wamberg, and is co-founder of Genome Medical, outside the submitted work.

### Funding Statement

Funding support was partly provided by grant 5R01HL143295 from the National Heart, Lung, and Blood Institute (LLV, CLZ, RCG, HLR, MSL).

### Author Declarations

This project has been reviewed and approved by the Mass General Brigham IRB.

