## Supplemental Materials for "A Framework for Automated Gene Selection in Genomic Screening"

#### SUPPLEMENTARY MATERIALS

**Supplementary Methods**

**Supplementary Figure Legend**

**Supplementary References**

**Supplementary Figure 1**

**Supplementary Tables 1-2**

#### Supplementary Methods

##### Selection and Parsing of Gene-Disease Association Resources

The Human Gene Mutation Database (HGMD, v2020.2) included 10,558 genes with at least 1 classified variant and was parsed to maintain classification and latest publication date associated with each variant[1]. Online Mendelian Inheritance in Man (OMIM) Morbid Map (downloaded August 12, 2020) was limited to the 16,895 genes with Entrez Gene IDs[2]. The ClinVar variant summary file (variant_summary.txt.gz; downloaded August 12, 2020) consisted of 11,196 genes with Entrez Gene IDs. After excluding copy number variants spanning multiple genes, the file was parsed to only include classification status, last evaluated date, and annotations for 2-star submissions for all variants[3]. The DisGeNET RDF dataset (all_gene_disease_associations.tzv.gz; v7.0) included gene-disease association (GDA) scores for associations between 21,671 genes and 30,170 diseases[4-6].

##### Data Sources Used for Validation of Diagnostic and Screening Gene Lists

The following manually curated gene-disease association resources were used for evaluation: 1) 1,490 GDAs evaluated in MedSeq[7]; 2) 1,514 GDAs evaluated in BabySeq[8]; 3) 1,043 gene curations in 824 genes captured by ClinGen as of August 12, 2020[9]; and 4) 232 and 306 panels from the PanelApp Australia and Genomics England PanelApp[10]. **Supplementary Table 1** lists the classification schemes for each of these resources and how they map to each other for evaluation purposes. The classification grouped as "Other" refers to GDAs classified as conflicting, refuted, disputed, no reported evidence, trait, pharmacogenomic association, only claim is from GWAS, and does not meet criteria. We assigned genes with classifications for multiple different diseases the strongest classification in each dataset.

For the crowdsourced PanelApp data, data was accessed via their APIs on July 23, 2020. We intersected classifications between PanelApp Australia and Genomics England PanelApp to minimize the effect of less well-curated classifications. The 4 panels from the Genomics England PanelApp listed as research panels were removed from analysis.

Discordant classifications between PanelApp England and Australia were excluded from further analysis.

**Genome Sequencing**

Genome sequencing was conducted at the Broad Institute of MIT and Harvard in 2019 and 2020. Genome sequence was generated from genomic DNA that is fragmented and barcoded followed by sequencing on the Illumina NovaSeq instrument with a minimum 20X coverage for 95% of all non-N positions in the genome. Reads are aligned to GRCh37 using the Burrows-Wheeler Aligner 0.7.15 (BWA-mem)[11,12], and variant calls are made using the Genomic Analysis Tool Kit HaplotypeCaller 3.5 (GATK)[13, 14].

Variants in genes of interest were subsequently filtered to identify: (1) variants classified as DM in HGMD or P/LP in ClinVar with a minor allele frequency ≤5.0% for all subpopulations in the Genome Aggregation Database v2 (gnomADv2)[15]; (2) nonsense, frameshift, start-loss, and +/-1,2 splice-site variants with a minor allele frequency ≤1.0% for all subpopulations in gnomADv2, or (3) variants previously classified by our laboratory as P/LP. The evidence for phenotype-causality was evaluated for each variant identified from the filtering strategies listed above and variants were classified based on the 2015 American College of Medical Genetics and Genomics/Association of Molecular Pathology guidelines[16] with ClinGen rule specifications (https://www.clinicalgenome.org/working-groups/sequence-variant-interpretation/).

**Supplementary Figure Legend**

**Figure S1:**

(A) Concentric pie chart of variants identified in the diagnostic (outer ring) and screening (inner ring) gene lists among 45 healthy genomes. (B) Graph of number of variants identified from the diagnostic and screening gene lists, and the number of reportable variants per genome.

### Figure S1

**A**

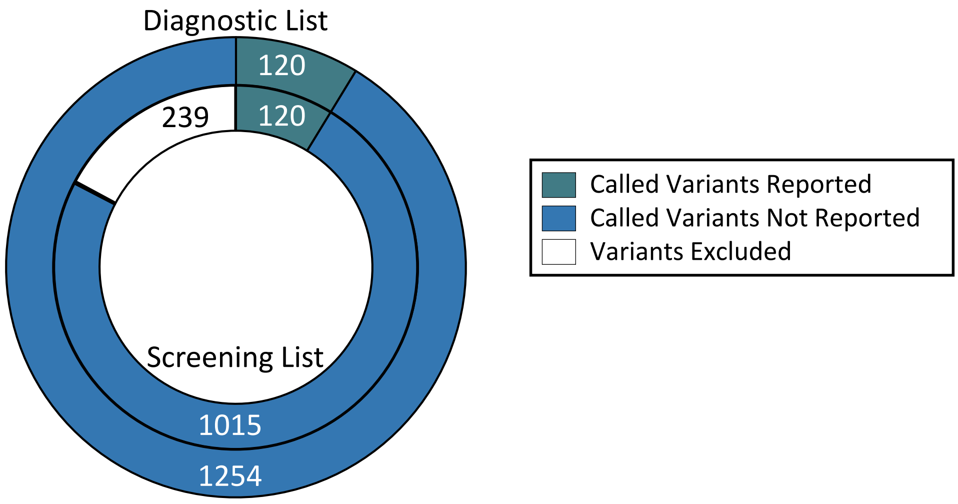

Diagnostic List
120
120
239
Screening List
1015
1254
Called Variants Reported
Called Variants Not Reported
Variants Excluded


**B**

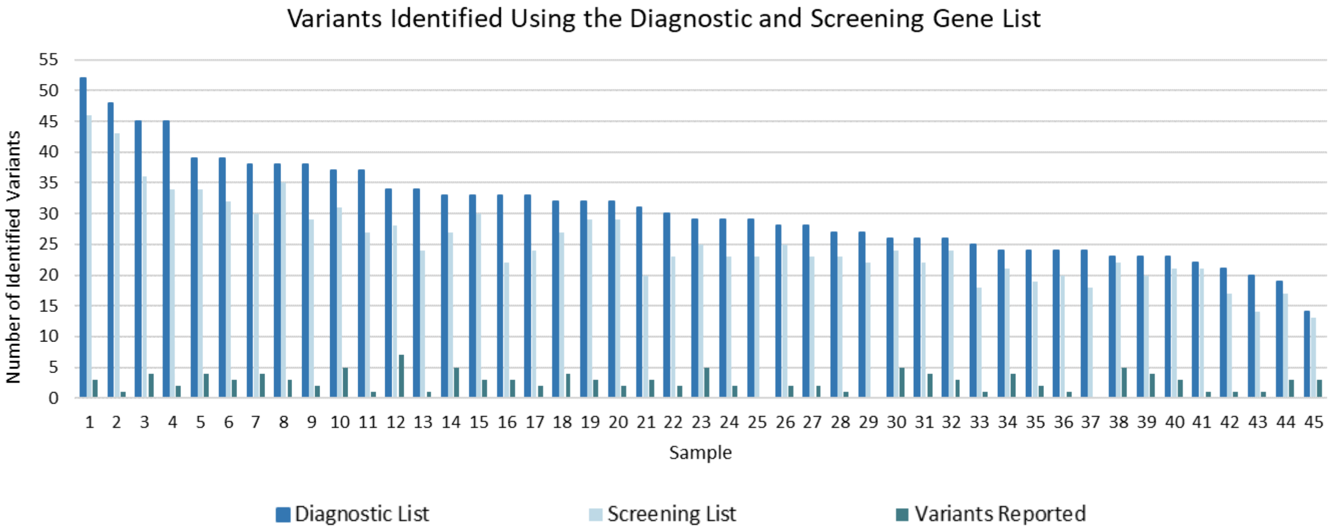

Variants Identified Using the Diagnostic and Screening Gene List
Number of Identified Variants
Sample
Diagnostic List
Screening List
Variants Reported

**Supplementary Tables**

##### Table S1: Overall strength of the classification assigned to each gene-disease association by each dataset.

| Strength of supporting evidence | Gene List | | Gene-disease association classifications used in each dataset | | | | |
|---|---|---|---|---|---|---|---|
| | | | Genomes Analyzed | MedSeq Project | BabySeq Study | ClinGen | PanelApp |
| Strong | Diagnostic Gene List | Screening Gene List | Definitive | Definitive/Strong | Definitive | Definitive | Strong (Green) |
| | | | Strong | | Strong | Strong | |
| Reduced | | | Moderate | Moderate | Moderate | Moderate | Moderate (Amber) |
| | | | Limited | Insufficient | Limited | Limited | Not Enough Evidence (Red) |
| Other | | | | DNMC; only claim is from GWAS; PGx association only; Trait | Conflicting | No Reported Evidence, Refuted, Disputed | |

DNMC: Does Not Meet Criteria; GWAS: Genome-Wide Association Study; PGx: PharmacoGenomics; Trait- gene only associated to a benign trait or to a biochemical finding without a clinical phenotype

##### Table S2: Diagnostic and Screening Gene List

Available separately

LINK: https://docs.google.com/spreadsheets/d/1VabFne_4TqEHxczwurwC8gokE0Q9fXCfbLlS5-9SqYs/edit#gid=1463456679
